# Understanding the barriers and facilitators to delivering peer support effectively in England: a qualitative interview study

**DOI:** 10.1101/2025.01.14.25320189

**Authors:** Una Foye, Natasha Lyons, Prisha Shah, Lizzie Mitchell, Karen Machin, Beverley Chipp, Stephen Jeffreys, Tamar Jeynes, Karen Persaud, Vicky Nicholls, Ruth E Cooper, Andrew Grundy, Tamara Pemovska, Nafiso Ahmed, Rebecca Appleton, Julie Repper, Sonia Johnson, Brynmor Lloyd-Evans, Alan Simpson

## Abstract

**Background:** Peer support roles within mental health services are rapidly increasing in number and scope in the UK and internationally. This paper explores the facilitators and barriers to delivering peer support effectively, as experienced by Peer Support Workers (PSWs) in a range of services and settings.

**Methods:** We conducted semi-structured qualitative interviews with paid mental health PSWs working across a range of settings in England. We took a collaborative, participatory approach. Interviews were carried out by researchers with experience of living with and/or supporting others with mental health conditions, and for some having experience delivering peer support themselves, and data were analysed using methods guided by general principles of thematic analysis.

**Results:** We interviewed 35 PSWs. Their roles were varied, spanning a range of mental health areas, and regions, but overarching facilitators and barriers were identified that appeared important across roles and settings. These included the need for roles to have flexibility with some structure and boundaries; the need for support, supervision and training to ensure PSWs are skilled in delivering the unique elements of their job; the importance of working with a strong team and leaders who support and value PSWs to help navigate the challenging personal aspects of being a PSW; the complexity of working in mental health systems where systemic factors such as funding, pay and progression can have a considerable effect both on the personal experience of PSWs and on the value placed on PSWs in the wider organisation.

**Conclusion:** Our findings highlight the complexity of PSWs, with a wide variety both of facilitators allowing them to carry out roles effectively and barriers to doing so. As PSWs numbers and the scope of their roles increase, awareness of barriers and facilitators needs to inform PSW job roles, support systems and integration into teams and mental health systems.

## 1. Introduction

Peer support is an organic and naturally occurring experience that has become formalised and disseminated within mental health services in recent decades (1,2). Within mental health settings peer support has been defined as a specialised approach in which someone with lived experience of a mental health conditions supports someone else who is also experiencing mental health difficulties (1). This approach tends to place greater emphasis on person-centred outcomes, such as social inclusion and empowerment, rather than traditional clinical outcomes, such as psychiatric symptomatology (3). By providing emotional, social, and practical support, peer support workers (PSWs) aim to achieve improvements in a range of health and wellbeing outcomes, including building people’s knowledge, skills and confidence to manage their condition and improving quality of life and social functioning (4). The value of embedding lived experience in mental health services is recognised internationally, with peer support recommended and implemented countries including Australia, Canada and the USA (5). In the UK, the implementation of peer support has grown across a range of mental health services, including Voluntary, Community and Social Enterprise (VCSE) sectors and NHS services (6).

As peer support develops internationally, an understanding of barriers and facilitators to delivering it effectively is needed to allow design of roles, support systems and integration into teams and systems in a range of contexts. Our recent umbrella review (5) summarised existing evidence on facilitators and barriers to implementing and delivering peer support within mental health services were explored. We found that factors promoting successful implementation included adequate training and supervision, a recovery-oriented workplace, strong leadership, and a supportive and trusting workplace culture with effective collaboration. Conversely, barriers included lack of time, resources and funding, and lack of recognised PSW certification.

In England, peer support has grown in the past decade a recent workforce stocktake identified more than 800 PSWs employed across 34 mental health NHS providers in England (62% of the 55 mental health Trusts in England at the time) services nationwide England, with approximately 86% directly employed by the NHS and the remaining 14% employed by external parties – for example, organisations within the voluntary sector (7,8). Currently, implementation plans and policies project a further increase in the peer support workforce in England, and the development of roles within a more diverse range of services than ever before, e.g., outside statutory services, within specialist mental health settings such as eating disorder services, and at managerial levels (8)The widespread introduction of peer support roles provides an opportunity to explore the experiences of this diverse and growing workforce to ensure that we learn from the factors that facilitate and impede them carrying out their roles effectively. Our study aims to explore these across among PSWs working in England.

## 2. Methods

This study was conducted by the NIHR Policy Research Unit in Mental Health (MHPRU), based at King’s College London (KCL) and University College London (UCL), which delivers evidence to inform government and NHS policy in England, agreeing a programme of rapid research with policymakers.

We took a collaborative, participatory approach to conducting qualitative interview research (9,10). The study is reported according to the COREQ checklist (Consolidated Criteria for Reporting Qualitative Research) (see S1 Appendix) (11). The study was approved by the UCL (University College London) Research Ethics Committee (REF: 19711/001, obtained 9^th^ January 2023).

### 2.1. Research team and positionality

The research team consisted of academic researchers and lived experience researchers, who have personal experience of using mental health services or supporting those who do, from the MHPRU. A project working group was established which met regularly throughout the project, consisting of the research team and key stakeholders (e.g. people working in the VCFSE peer support sector).

Interviews were carried out by lived experience researchers (TJ, KM, BC, SJ, PS, LM, KP, VN, NL), with some of these LERs also having experience working in peer support, including of working as PSWs in mental health services and training peer workers. Interviews were conducted independently by the LERs, with most accompanied by a university-employed researcher who was responsible for the interview recording (UF). LERs received training in interviewing skills and data analysis. Following the interviews, LERs could debrief with a member of research staff if needed or take part in regular reflective spaces to discuss their experiences of conducting the interviews. A team of LERs and other members of the research team coded the data and were actively involved in the analysis and writing of findings.

Within peer support work, a range of terminology describes providers and recipients of peer support. Within this paper we aim to use consistent language where possible when referring to peer support workers (PSWs) and those they support, e.g., service users, but acknowledge that there is a wide range of terms that are used within the peer support literature and by the participants involved within this study.

### 2.2. Participants

Participants were eligible if they were adults (18+ years), who were employed to use their personal experience of mental health conditions in paid PSW roles across a range of mental health services (e.g. NHS, VCFSE). We excluded people: working as volunteer PSWs; working in services solely supporting people who experience organic neurological conditions or substance misuse, or who solely provide peer support through online forums or via online psychoeducation or psychological interventions.

We recruited using purposive sampling to ensure representation of a range of PSW roles, and age groups. Recruitment occurred through circulating the study advert via i) organisations that provide training for peer support, including organisations who support people who may be underrepresented in peer support research, such as people working with children and young people or with minority ethnic groups; ii) the national and local service user networks known to members of the study team; iii) social media. Interested participants contacted the research team via email, who confirmed eligibility, provided the participant information sheet and offered a phone or video call to discuss the study. Participants gave informed consent to participate in the study. Participants were offered the option of an in person or remote interview by video or telephone.

### 2.3. Data collection

To develop the topic guide, we held an initial meeting as a research team to discuss potential content. An open document was then shared with all lived experience researchers to add specific topics and questions that would be important to include, guided by experiential expertise, knowledge of peer support practice and current gaps in the literature. We then developed the topic guide through an iterative process of development with regular meetings with the project working group and research team. The topic guide was piloted on the first 10 interviews (See S2 Appendix). We then held a sense checking workshop with the project working group was to discuss initial findings and agree and make minor changes to the topic guide. The topic guide explored the following areas with PSWs:

- Their roles and unique aspects of their professions.
- The support they provide.
- Their ability to use their lived experience as they would choose to.
- Barriers and facilitators they experience.
- Experiences within mental health teams.
- Supervision, training and career progression.
- Their impact on the mental health system and wider community.

All interviews took place between March and August 2023, remotely via video call on Teams. and were audio-recorded. Before the start of the interview, informed consent was obtained and demographics collected from participants. Participants were informed that their interviewer was a lived experience researcher. Interviews ranged from 30 minutes to 2 hours with the average interview length being 1 hour 14 minutes. Participants were emailed a study debriefing sheet after the interview (giving a list of resources and the option to speak with one of the academic researchers for support, should they feel any distress after the interview). Participants received a gift voucher (£20) for taking part.

### 2.4. Data analysis

Interviews were recorded, transcribed and anonymised prior to coding. Data were analysed by the research team (lived experience researchers and academic researchers) using collaborative methods refined during previous studies conducted by the MHPRU team (9,10). The analysis process used methods guided by general principles of thematic analysis (12). The analysis occurred in 5 stages:

1. **Preliminary coding:** the research team (lived experience researchers, n=8, academic researchers, n=2) each coded one transcript of an interview (using the same transcript). Using a coding matrix, they noted thematic ideas developed and interpreted from the transcript that seemed to articulate core aspects of the interviewee’s experiences and to fit within the three research question domains.
2. **Developing a coding framework:** the research team met to review and combine matrices. Similar thematic ideas were grouped together to develop a provisional coding framework. The provisional coding framework was then circulated to the research team and a meeting held to review and refine the framework. A smaller research team (lived experience researchers, n=4, academic researchers, n=1) piloted the framework on one-or-two different interviews each (in NVIVO (version 14), Excel or Word; (13–15). Another meeting was held to review and refine the framework after piloting, with decisions made about dividing or adding new themes. An initial coding framework was thus developed.
3. **Initial coding of data:** the smaller research team coded all interviews from transcripts using the coding framework (in NVIVO or Microsoft Word; (14,15)).
4. **Refining the coding framework:** throughout interview coding the smaller research team met regularly to discuss whether the interviews they coded offered a good fit with the framework or to agree on revisions to the framework. A revised framework incorporating any adapted or new themes was then produced.
5. **Writing up the analysis:** The revised framework was used by the smaller research team to write up each theme, using example quotes and annotations to write an analytical narrative around the data. This involved regular meetings to i) read, review and further consolidate the themes; ii) draft the description of the themes; iii) review and finalise the theme write-up. We made further refinements to the themes through discussion with the wider working group.

To help address the research aim of understanding the wide-ranging barriers and facilitators experienced by PSWs across the system, themes were organised into wide categories that followed Bronfenbrenner’s ecological theory (16). Key themes were therefore organised across the three levels of the macro, meso and micro systems to provide a framework to help understand the complexity and interconnectivity of the issues being explored and to allow an understanding of the research questions from the systematic/ organisational perspective, local on the ground perspective, and the individual perspective.

## 3. Findings

We conducted interviews with 35 eligible participants. Despite confirming eligibility prior to the interview, four participants were excluded post-interview as it emerged during the interview that they did not meet eligibility criteria for their peer support role, e.g. they were working in unpaid roles. No participants withdrew from the study after the interview. Participant characteristics are shown in Table 1. Most participants were from a white British ethnic background (n=23), were aged between 35-44 years (n=11), and female (n=23). The majority of participants worked in NHS settings (n=21), with fourteen working in other settings such as charities or community-based groups. Considerable variety in job title was found among participants, 11 identified their job as “peer support worker”, and 12 identified as senior peer support roles or peer support coordinator roles. Other job titles included peer consultant, peer coach, peer specialist, peer leader, and peer recovery worker.

**Table 1:** Participant characteristics.

**Table 2:** Key facilitators and barriers to delivering effective peer support identified by system level.

Themes are presented using three levels representing themes coded to the micro (personal), meso (organisational) and macro (systemic/ societal) levels. Each theme level has a number of subthemes outlined in figure one. Figure 1 highlights the key facilitators and barriers to delivering peer support associated to these domains that are further discussed and exemplified in the following section. Data extracts and quotations to illustrate each theme and key analytic points are presented in a supplementary table (S3 Appendix).

**Figure 1:**
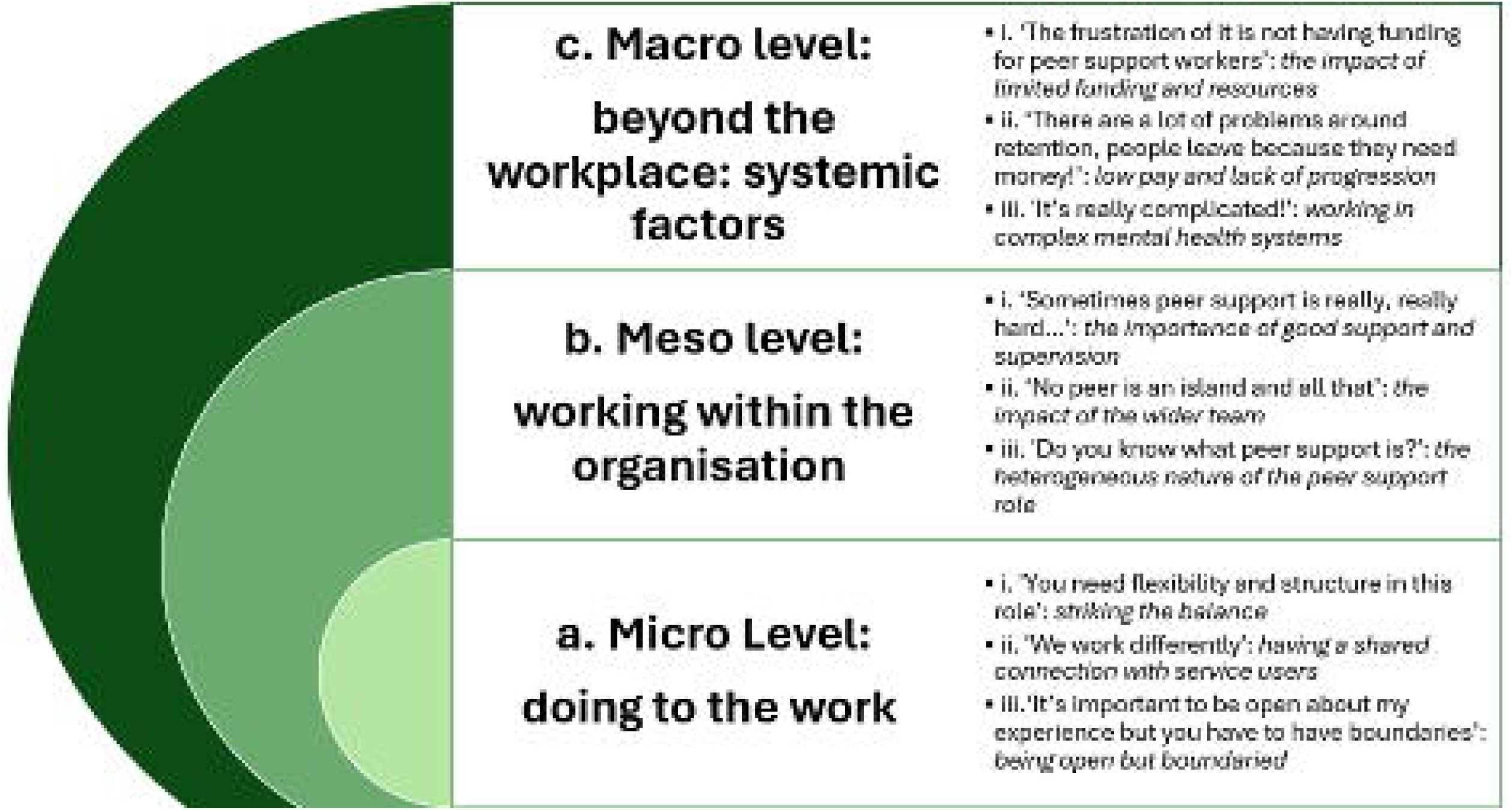
Key Themes.

### a. Micro level: doing the work

Participants spoke about the core values and unique elements that make the peer support role different to that of their non-lived experience colleagues. When exploring elements of what PSWs do at an individual level of the role, there were three core areas in which influences on how effectively peer support could be delivered were identified; i) exploration of the need to have flexibility in the work but within supportive structures), ii) the unique element of using their lived experience to be an bridge between clinical colleagues and service users, and iii) the personal qualities considerations needed to deliver this (e.g., the role of boundaries and self-awareness). These three subthemes are explored below:

**i. ‘You need flexibility and structure in this role’: striking the balance**

Many participants spoke about the need for the role to be flexible in its nature to ensure that they are delivering a person-centred approach, a core value of peer support work. Peer workers described a tension between flexibility that can be useful in developing the role in a way that feels right to them and the service users and working with a structure that provides clarity and security, which prevents being asked to undertake work that does not seem a good fit with a peer role. While flexibility was seen as an important facilitator to delivering the core elements of the work, it was also felt to be a barrier to delivering effective peer support for some participants, particularly for those working in third sector roles where it was felt that there was too much flexibility and that there was a need for some structure in the role to aid clearer expectations and boundaries for the role. It was felt that having clearer structure and expectations would facilitate PSWs to undertake their role more effectively as it helped to ensure the role was contained and understood. Without this, some participants felt their role was ambiguous, evidenced by people asking them to fulfil tasks outside of their remit, e.g. helping with physical restraints on inpatient wards. In contrast, participants who worked in more structured roles, i.e., roles with fixed guidelines regarding types of support offered (e.g., delivery of structured self-management tools) and the number of sessions that can be offered, noted that this acted as a barrier to them delivering peer support as intended as they were restricted in what they could offer. Furthermore, there were concerns from peer workers that the role was becoming too closely integrated into standard clinical practice.

**ii. ‘We work differently’: having a shared connection with service users’**

At the core of the work delivered, PSWs felt it was important that they kept a clear distinction between themselves as peers and clinical staff, with the shared experience as a core element that helped to distinguish this difference in how individuals connect and experience the support provided by a peer worker in contrast to a clinician. This shared experience was seen as a key factor in facilitating a connection with service users and building the relationships with them. Examples of peer workers being able to connect with service users who are either disengaged from or perhaps neglected by traditional clinical services displays how a shared connection can be used to improve care for service users and help them re-connect with services. Peer workers therefore have unique skills which they report enable them to build connections between clinical services and service users that is embraced by professionals and service users alike.

This shared experience between service users and peers was felt to be an important facilitator to delivering effective peer support, reducing power dynamics and hierarchies and creating more of an equal playing field. This is particularly felt to be important where people have had poor experiences of mental health services, have experienced iatrogenic harm, or have mistrust in services due to cultural or systemic barriers. Through a PSW being able to understand the impact of these harms on the service user, it creates mutuality and helps them feel the peer is there to support them and their goals, instead of operating as a component of the system. Furthermore, peer workers felt this shared experience moved beyond experiences of mental illness or problems to encompass a more diverse, intersectional lens, e.g., understanding not only sharing an experience of mental health conditions but also understanding how this may be experienced from the perspective of a man, or someone from a black background. While there was real value felt by having peers sharing themselves, some peer workers from ethnic minority backgrounds faced challenges when they were working in wider teams with no colleagues from similar backgrounds or when they experienced initial prejudice from service users during their work.

**iii. ‘It’s important to be open about my experience but you have to have boundaries’: being open but boundaried**

A core feature of peer support work is the PSW and service user having lived experience and shared understanding of mental ill health, but participants reported that to do this in a healthy, safe and effective way they needed to be clear about how much of themselves and their experiences they are willing to share with others. Sticking to such boundaries was considered important in avoiding burnout and relapse, as participants said that the emotional burden of sharing their own traumas and reliving difficult experiences, as well as hearing other people’s pain was challenging and not always acknowledged by colleagues and managers. This was also enhanced by having clear personal boundaries about how much you were willing to share and having self-awareness of your own triggers.

Participants felt that having training in how to utilise lived experience, set boundaries and have self-awareness about sharing was seen as key in peer support roles to ensure safety of self and service users. A further facilitator to ensure PSWs could work within safe boundaries was identified as having a clear job role, and clear expectations and prescribed limitations. Where participants faced job ambiguity, there was a sense of being unsure of the limits that they could set in their work, and perhaps being subject to pressure from their clients, or co-option by staff to assist with other work.

### b. Meso level: working within the organisation

Participants recognised that they did not deliver their work in a vacuum, and that it was important to contextualise this personal delivery of work within the wider working culture in which they were situated. The support they received from managers and colleagues, as well as the connections they had with other team members all had an impact on how they could deliver their work. Furthermore, how peer support work was viewed, whether it was valued, and the knowledge about PSW across team members and wider colleagues was felt to influence this. These themes are explored in more detail below:

**i. ‘Sometimes peer support is really, really hard…’: the importance of good support and supervision**

Given the emotional responsibility and intensity that is experienced by PSWs, participants felt good support was necessary to be able to deliver effective peer work. Most participants reported feeling well supported in their role, with high value placed on the support they received from their managers. Several peer workers in the third sector reported more inconsistency in or absence of the support element that their NHS counterparts reported.

It was felt that this support should be flexible to mirror the nature of the PSW role, with PSWs reporting the benefit of utilising a range of support structures including from direct line managers as well as through external clinical supervision with lived experience practitioners and other reflective spaces. Having shared ownership of the agenda for formal supervisions and support sessions was seen as helpful to many participants as it allowed them to utilise these spaces for what was important to them and their role. This meant elements of the role that might cause anxiety and the finer details of working with service users could be discussed and supported. While formal sessions were valued and necessary to help peers do their job well and safely, informal support was also helpful, including managers being contactable outside of pre-arranged meeting times. This informal support was particularly valuable when things were difficult, or for people predominantly working alone, e.g., doing lone work, or work in rural areas.

**ii. ‘No peer is an island and all that’: the impact of the wider team**

Most participants who worked within a wider, multi-disciplinary team felt that they had good working relationships with their coworkers within their immediate team and that this contributed to their enjoyment of the role. In addition, these positive team relationships were felt to add to the informal support that peer workers received. For those who were in large multidisciplinary teams (MDT), being integrated into a team with people in different roles was beneficial, as it allowed them to learn from others and provided safe spaces to discuss issues in their work and problem solve. This was particularly felt across NHS roles. For those who were able to connect with other peer workers this was felt to enhance their experience of being a PSW as the shared experience of doing the role built a connection and helped them to share knowledge and skills to improve their practice. For some without these connections, they reported feeling isolated in their role.

While most participants felt they had positive working relationships with colleagues in their team some faced challenges when being open about their experience of living with mental health problems. Specifically, they noted the negative impact of colleagues’ assumptions about those with lived experience and fears regarding an individual’s vulnerability or risks, and concerns about saying the wrong thing were felt to impact working relationships and work more widely. This feeling of fear of colleagues who are clinicians was more prominent in areas where peer work was new, e.g., eating disorder services. Training team members who are not peer workers in peer support principles was seen as beneficial to the wider team, helping to break down barriers to promote acceptance and integration of the peer role.

**iii. ‘Do you know what peer support is?’: the heterogeneous nature of the peer support role**

The peer support role was felt by participants to be heterogeneous and diverse, with many regional and local variations, reflective of the wide and varying nature of mental health services. This contributed to a lack of clarity regarding the roles, e.g., the role is hard to define and understand. Lack of understanding of peer support roles had an impact on practical day-to-day working, with participants being asked to undertake tasks outside of their role and receiving unsuitable referrals from colleagues. More importantly, participants felt that the lack of understanding concerning peer support roles reflected the lack of value being placed on peer support work more widely. This ambiguity was more pronounced in services where peer support work was new in the organisation, compared to where peer support was established.

Facilitators that were felt to help address these issues included having clear strategies, protocols and systems in place, preferably co-designed with service users, and to have peer support visibility, for example through staff training or representation at higher levels within leadership teams. In addition, having managerial and leadership support was felt to facilitate the development of such policies and strategies to embed these roles. Having suitable peer workers on executive boards and governing bodies or developing more lived experience or peer roles at higher bands were all felt to improve visibility, staff buy-in and patient acceptance of peer support while also increasing the understanding and value of peer work across managers and staff. For some, this increased ‘professionalisation’ of PSW as a role was seen as a positive factor as it addresses this issue of role ambiguity and by having recognised qualifications this may in turn increase the value held of these roles. For others however, this was seen as a barrier to the core values of PSW, as it works towards removing flexibility from the role and assimilating it into the clinical system.

### c. Macro level: beyond the workplace – systemic factors

There was acknowledgement among participants of wider systemic issues affecting PSWs such as funding, pay, job instability and career progression. It was important to consider how these wider, structural factors have an impact on the delivery of peer support work, as well as their personal impact on the PSWs. These themes are explored in more detail below:

**i. ‘The frustration of it is not having funding for peer support workers’: the impact of limited funding and resources**

Almost every participant noted frustrations or challenges facing peer support work because of limited funding and a lack of resources both for the current workers and for the further development and growth of peer work set out in key policies and strategies. Frustrations included the stagnant nature of the role due to limited resources for development, training or progression, as well as the challenging interface this creates when informing service users there are not resources for longer term support. The lack of resources not only referred to limited funding for staffing or support systems but also for more basic elements needed to do the job. Participants reported not having access to computer equipment, funds to pay for travel to attend vital training sessions, a lack of office space which for one participant lead to them having to do all meetings including supervision and client meetings in public coffee shops, and not being able to claim expenses. Such restrictions around allocation of funding, resources or expenses had an immediate impact on people’s ability to deliver their work.

Funding decisions was felt by participants to be related to the value placed on PSW. While it was acknowledged that services for mental health are underfunded and stretched, it was felt that this was more prominent for PSW with a lack of funding for long-term posts and lack of investment into resources like training being felt strongly. While not unique to PSW, the fact these issues were perceived to be hitting PSW harder than other workers suggested that peer work was not as valued as clinical roles in the organisation.

**ii. ‘There are a lot of problems around retention, people leave because they need money!’: low pay and lack of progression**

Overall, the low level of pay and lack of progression for PSWs was felt to be a considerable issue facing this area of work. Low pay was felt to represent the lack of value placed on PSW in the mental health system, with some participants feeling that it showed that the importance of peer work still wasn’t recognised for being the vital but hard and effective work that it is. Other factors related to how peer support was organised in practice that added to participant confusion and frustration regarding pay and progression for these roles included inconsistent pay scales and bandings, and the set-up of these roles, e.g., some staff being paid by a third sector organisation but embedded in the NHS. The ambiguity about bandings, career development, and lack of transparency in the role descriptions creates a very unclear picture of peer support and its place in the wider system. This was not consistent across participants, with some people acknowledging that they are happy with their banding or pay scales. Participants were motivated to continue this work due to their love of the role and its role in their recovery.

In addition to pay, there were challenges regarding progression in the role. Many participants felt that although they loved the nature of peer work, the lack of progression and career development in the role may mean they have to look to other employers or other roles to fulfil their career needs. It was suggested that many people leave the role due to the lack of career prospects and consequently, the organisation loses the PSWs’ lived experience expertise. Some participants reported they were happy with their current role and did not want to transition into a different role to develop their career.

**iii. ‘It’s really complicated!”: Working in complex mental health systems**

Working within the wider mental health system brought complexity to PSW roles. This was more pronounced for those working in the NHS as this has its own politics, language and structures to navigate. Working within multidisciplinary teams, where there were many competing demands and views to negotiate and understand could be challenging. Being embedded in teams and collaborating with a range of services was felt to help peer workers understand how services may link up and to see the bigger picture but was still experienced as very challenging, frustrating, and ever changing. As well as the nature of the NHS as a very large organisation with many moving parts, participants noted the challenge of having to navigate the siloed working and red tape within the system in which there was felt to be a lack of cohesion between and within services. As a result, some feared that peer work would itself become overtaken by this culture rather than retaining its core elements as a flexible and person-centred way of working. Some participants reported that they felt hopeless in navigating a complex system which contributed to feeling burnt out and challenged in delivering interventions that instil hope to clients. This was particularly salient for those working in rural areas in which they were often in a small team, or where there was only one peer worker.

## Discussion

The findings presented within this study provide insight into the facilitators and barriers that influence whether PSW are able to deliver peer support effectively. While the findings identify a varied workforce and experiences, with PSWs spanning a range of mental health settings, specialisms, and regions, there were overarching facilitators and barriers identified as important to PSWs. These included the need for support, supervision and training to ensure PSWs are skilled in delivering the unique elements of their job, e.g., understanding boundaries for working and sharing their lived experience. This not only highlights the need for self-awareness and personal strategies for engaging with the work, e.g., adaptability and flexibility to deliver person-centred care, but also allows us to view the important role of team members, managers and organisations in providing space and support to allow PSWs to do this work. Organisations must take their duty of care towards PSWs seriously and ensure clear guidelines are in place regarding engagement with clients and the importance of self-care is regularly addressed.

The findings from this paper show that team preparation and ongoing support for teams is necessary; this is not only a responsibility for managers and supervisors but also a role for co-workers and wider team members as this helps to foster a positive working culture in which team members can learn from PSWs, and PSWs can feel supported in the challenges of the work, drawing on and sharing their personal experiences of mental distress and recovery. It was recognised that to enable these key factors there is a need for wider systemic supports and cultures, including having clear PSW strategies to ensure clarity about what PSW is, and what the role adds to the current systems and for whom. Furthermore, there is a need for sustainable funding, with clear opportunities for PSWs to develop and grow, with clear career pathways. These systemic elements all add to a culture of value for PSW, and work towards tackling key barriers observed across all levels, e.g., motivation, ambiguity of role, and flexibility in the role.

Many of these issues confirm and validate previous reported findings (2,5,17–20). However, by providing insight into the layers at which these barriers and facilitators exist and inter-relate, and by helping to understand how PSWs can be supported personally and at an organisational and systemic level, we can help inform how workforce plans to increase PSW roles in mental healthcare are implemented effectively (7,8,21–23). Many of the concerns identified in this study are addressed within these frameworks and policies, therefore future work needs to explore whether these guidelines are being implemented and whether concerns about pay and career development are being fully addressed.

One key element highlighted within these findings was the ambiguity of PSW roles and what is considered peer support. This is a longstanding issue that continues to face PSWs globally (24–26), and is particularly important as PSW roles keep developing to include the evolving nature of remote and teleworking, as well as the expansion of PSWs into specialist areas such as services for eating disorders or rough sleeping. As a result, systems are needed to ensure the role can work in this way, through provision of training to the PSWs and their colleagues, recognised qualifications, and the provision of suitable supervision and support.

In addition, the findings of this paper add to our understanding of the potential friction between the PSW role as a distinct and independent role that is positioned to be an ally for service users within clinical settings, and the need or want for PSWs to be more integrated into wider clinical teams (27). There is a real need to strike a balance in which the role can continue to provide the allyship and flexibility that is unique to the PSW role, without losing the closer working and support that can be provided by clinical colleagues and within the wider organisational systems. There was recognition that the system has a place for both elements, although a distinction was needed to ensure that the unique element of PSW utilising lived experience perspectives was enabled and not lost or sidelined.

### Strengths and limitations

This paper has a strength in the wide range of participants that were included, showing the diversity and range of peer support roles in England. This allows our findings to exemplify the commonality of many of the facilitators and barriers facing the whole workforce. Furthermore, this study used a self-selection recruitment approach and only included views from individuals currently employed within paid PSW roles, and many of these had only been in position a relatively short time. Future research would benefit from including PSWs who have both been employed longer, and those who have left the profession to understand the longer-term impact that this work may have on them, and how the barriers identified within this work may impact recruitment and retention. n.

### Implications for policy and practice

The findings in this paper have clear implications for both policy and practice. At a workforce management level these finding present indicators for the needs of the PSW workforce by identifying the factors that help and hinder them doing their roles. There is a clear need for attention to be paid to the career development pathways aligning with the progression pathways that exist for other healthcare professions. As noted within this paper, without these clear progression pathways skilled PSWs are likely to leave for other roles such as nursing or psychology where there are clear pathways for promotion and growth. These issues can only be done when there are longer term secure funding streams in place to tackle the key issues of fixed term contracts and job insecurity within the field. Some participants had started peer work as volunteers. There is a place for this, and it can help someone decide if it is a potential career for themselves or not without the commitment. However, in mental health care in particular, stability of staff and those who support you is important, and volunteer roles are likely to be less stable than employed PSWs. Finally, volunteers must not be used as a substitute for paid staff and PSW’s must not be used as a cheap option to dispense with clinically trained staff. In a healthy organisation all these will work together in a symbiotic way.

In practice, the emotional burden of this work creates a need for more support for PSWs at the individual level through the provision of supervision and clear line management, and support from other peers and clinical colleagues. While training is growing in the field and is highly valued by PSWs, these findings demonstrate that there is a need to provide additional training and awareness of peer support across teams including clinical colleagues and managers in leadership or senior organisational roles, something that is becoming part of the new normal in mental health services (28). Such training can ensure that the value of PSWs and clarity about the role, understood and valued in line with the regional and local policies and frameworks (7). Health professional educators also need to ensure that continuing, or continuous, professional development (CPD) and pre-registration training curricula include content on the nature and purpose of PSW roles in the modern mental health workplace (29). Furthermore, the findings indicate that organisations need a clear PSW strategy so that opportunities for development, promotion and influence are clear.

## Conclusion

In conclusion, this paper highlights the complexity facing PSWs noting numerous facilitators that can better support them in their roles and numerous challenges that can create barriers. Issues outlined note the importance of having clarity in the role; the need for balance between flexibility and structure in the role; the need for support in the form of connections with other lived experience workers, colleagues, and managers; as well as the need for recognition and value being placed on the PSW role through increased funding, promotion, qualifications and career development opportunities. While some of these issues have been previously identified within the literature, they remain significant to PSWs in todays’ workforce. Given the increased number of peer workers and a growing demand for their work and the increased reach that PSWs are having in mental health services, there is a real need for these barriers to be addressed and facilitators to be enhanced to ensure that peer workers can deliver their work effectively.

**Lived experience commentary (written independently of this study by Dr Hannah Lewis and Mark Holden, members of the wider MHRPU Lived Experience Researcher team).**

As lived experience researchers (LERs), we welcome this paper which explores the barriers and facilitators to implementing roles as peer support workers (PSWs), where people apply their experiential knowledge in supporting others to manage their mental health. It is encouraging to see a more “collaborative, participatory approach” when the research team conducts qualitative interviews with participants. However, it would’ve been better to also explore the positionality of the research team alongside similar demographics collected from research participants. The paper recognises how peer support is recovery-focused, and rooted in “person-centred outcomes, such as social inclusion and empowerment, rather than traditional clinical outcomes, such as psychiatric symptomatology”.

Furthermore, this paper alludes to tensions surrounding the mandate from the NHS to formalise PSW roles. It highlights the ambiguity in defining the role, which is a barrier for PSWs currently – and we share concerns. Whilst we appreciate that formalising these roles can act as a facilitator by enabling standardised career progression pathways and competencies, we acknowledge that this could appear to be contradictory to the grassroots origins of PSW roles. However, it is important to ensure that PSW roles have consistent, standardised training and good quality supervision applied across all mental health settings, enabling role-specific competencies and psychological safety for both PSWs and the people they support.

This paper is a positive step in considering some critical barriers and facilitators in embedding PSWs across mental health services. Whilst the benefits of their involvement in mental health services is undeniable – with their role “being a bridge” and fostering trust and hope – a PSW is only as good as the service they work in and should not be seen as a ‘silver bullet’ in mental health care. Hiring more PSWs should not detract from some of the systemic failings that remain in mental health service provision.

## Declarations

### Ethics approval and consent to participate

All procedures performed in studies involving human participants were in accordance with the ethical standards of the institutional and/or national research committee and with the 1964 Helsinki declaration and its later amendments or comparable ethical. Ethical approval was granted by the UCL (University College London) Research Ethics Committee (REF: 19711/001, obtained 9^th^ January 2023). All participants provided informed consent prior to enrolment in the study, including consent for publication of anonymised quotes.

### Consent for publication

Not Applicable

### Availability of Data

The data that support the findings of this study are available on request from the corresponding author. The data are not publicly available due to privacy or ethical restrictions.

### Author Contributions

All authors have read and approved the manuscript. Authors BLE and NL created the protocol for the study. UF conducted the recruitment for the study, and TJ, KM, BC, StJ, PS, LM, KP, VN, NL, TK conducted the interviews. UF, KM, BC, PS, LM, TK coded data, and contributed to the thematic analysis. All authors supported drafting and development of the manuscript.

## Acknowledgements

We would like to acknowledge the Mental Health Policy Research Unit team, Lived Experience Working Group members, and Project Working Group who have contributed and supported this work. Special thanks to Hannah Lewis and Mark Holden for their contribution to the paper by providing an independent lived experience commentary.

## References

1. Davidson L, Bellamy C, Guy K, Miller R. Peer support among persons with severe mental illnesses: a review of evidence and experience. World Psychiatry. 2012 Jun 12;11(2):123–8.

2. Repper J, Carter T. A review of the literature on peer support in mental health services. Journal of Mental Health. 2011 Aug 19;20(4):392–411.

3. Puschner B, Repper J, Mahlke C, Nixdorf R, Basangwa D, Nakku J, et al. Using Peer Support in Developing Empowering Mental Health Services (UPSIDES): Background, Rationale and Methodology. Ann Glob Health. 2019 Apr 5;85(1).

4. NHS England. Supported self-management: peer support guide [Internet]. 2017 [cited 2024 Jun 5]. Available from: https://www.england.nhs.uk/publication/community-capacity-and-peer-support/

5. Cooper RE, Saunders KRK, Greenburgh A, Shah P, Appleton R, Machin K, et al. The effectiveness, implementation, and experiences of peer support approaches for mental health: a systematic umbrella review. BMC Med. 2024 Feb 29;22(1):72.

6. da Costa M P, Foster R, Gillard S, Priebe S. Volunteering in mental health. In: Okpaku S, editor. Innovations in global mental health. Cham: Springer Internatinoal; 2019. p. 1–28.

7. Health Education England. National Workforce Stocktake of Mental Health Peer Support Workers in NHS Trusts. [Internet]. 2020 [cited 2024 Jun 5]. Available from: https://www.hee.nhs.uk/sites/default/files/documents/NHS%20Peer%20Support%20Worker%20

8. NHS England. Supported self-management: peer support guide. [Internet]. 2023 [cited 2024 Jun 5]. Available from: https://www.england.nhs.uk/long-read/peer-support/

9. Gillard S, Dare C, Hardy J, Nyikavaranda P, Rowan Olive R, Shah P, et al. Experiences of living with mental health problems during the COVID-19 pandemic in the UK: a coproduced, participatory qualitative interview study. Soc Psychiatry Psychiatr Epidemiol. 2021;56(8):1447–57.

10. Shah P, Hardy J, Birken M, Foye U, Rowan Olive R, Nyikavaranda P, et al. What has changed in the experiences of people with mental health problems during the COVID-19 pandemic: a coproduced, qualitative interview study. Soc Psychiatry Psychiatr Epidemiol. 2022;57(6):1291–303.

11. Tong A, Sainsbury P, Craig J. Consolidated criteria for reporting qualitative research (COREQ): a 32-item checklist for interviews and focus groups. International Journal for Quality in Health Care. 2007 Sep 16;19(6):349–57.

12. Braun V, Clarke V. Using thematic analysis in psychology. Qual Res Psychol. 2006 Jan;3(2):77–101.

13. Microsoft. Microsoft Excel. Microsoft Corporation; 2023.

14. Microsoft. Microsoft Word. Microsoft Corporation; 2023.

15. Lumivero. Nvivo Software v14. www.lumivero.com; 2023.

16. Bronfenbrenner U. The ecology of human development: Experiments by nature and design. Cambridge, MA: Harvard University Press.; 1979.

17. Kotera Y, Newby C, Charles A, Ng F, Watson E, Davidson L, et al. Typology of Mental Health Peer Support Work Components: Systematised Review and Expert Consultation. Int J Ment Health Addict. 2023 Aug 14;

18. Kuek JHL, Chua HC, Poremski D. Barriers and facilitators of peer support work in a large psychiatric hospital: a thematic analysis. Gen Psychiatr. 2021 May;34(3):e100521.

19. Burke EM, Pyle M, Machin K, Morrison AP. Providing mental health peer support 1: A Delphi study to develop consensus on the essential components, costs, benefits, barriers and facilitators. International Journal of Social Psychiatry. 2018 Dec 3;64(8):799–812.

20. Gillard SG, Edwards C, Gibson SL, Owen K, Wright C. Introducing peer worker roles into UK mental health service teams: a qualitative analysis of the organisational benefits and challenges. BMC Health Serv Res. 2013 Dec 24;13(1):188.

21. NHS. NHS Mental Health Implementation Plan 2019/20 – 2023-24 [Internet]. 2019 [cited 2024 Jun 5]. Available from: chrome-extension://efaidnbmnnnibpcajpcglclefindmkaj/ https://www.longtermplan.nhs.uk/wp-content/uploads/2019/07/nhs-mental-health-implementation-plan-2019-20-2023-24.pdf

22. Commonwealth of Australia. Fifth National Mental Health and Suicide Prevention Plan [Internet]. Canberra; 2021 [cited 2024 Jul 16]. Available from: https://www.mentalhealthcommission.gov.au/sites/default/files/2024-04/fifth-national-mental-health-and-suicide-prevention-plan-2021-progress-report-4.pdf

23. Gagne CA, Finch WL, Myrick KJ, Davis LM. Peer Workers in the Behavioral and Integrated Health Workforce: Opportunities and Future Directions. Am J Prev Med. 2018 Jun;54(6):S258–66.

24. Kemp V, Henderson AR. Challenges faced by mental health peer support workers: Peer support from the peer supporter’s point of view. Psychiatr Rehabil J. 2012;35(4):337–40.

25. Tseris E. The Expansion of the Peer Adviser Workforce: Opportunities and Challenges for Social Work. Australian Social Work. 2020 Apr 2;73(2):162–74.

26. Adams WE, Duquette R, de Wet A, Rogers ES. Competing allegiance in an unclear role: Peer and non-peer understanding of peer support in Massachusetts, United States. SSM - Mental Health. 2023 Dec;4:100245.

27. Sinclair A, Fernandes C, Gillieatt S, Mahboub L. Peer work in Australian mental health policy: What ‘problems’ are we solving and to what effect(s)? Disabil Soc. 2024 Aug 8;39(7):1656–81.

28. Jones N. Lived Experience Leadership in Peer Support Research as the New Normal. Psychiatric Services. 2022 Feb 1;73(2):125.

29. Charles A, Nixdorf R, Ibrahim N, Meir LG, Mpango RS, Ngakongwa F, et al. Initial Training for Mental Health Peer Support Workers: Systematized Review and International Delphi Consultation. JMIR Ment Health. 2021 May 27;8(5):e25528.

