## Supplementary material for "Understanding the barriers and facilitators to delivering peer support effectively in England: a qualitative interview study": Table 1

**Table 1:** *Participant characteristics*

| **Characteristic** | **Category** | **Number** |
| --- | --- | --- |
| Age (years) | 18-24 years | 6 |
|  | 25-34 years | 6 |
|  | 35-44 years | 11 |
|  | 45-54 years | 6 |
|  | 55-64 years | 5 |
|  | >65 years | 1 |
| Gender | Female | 23 |
|  | Male | 10 |
|  | Non-binary/gender self-defined | 2 |
| Ethnicity | White British | 23 |
|  | Asian or South Asian | 5 |
|  | Other white | 3 |
|  | Black Caribbean | 2 |
|  | Black African | 1 |
|  | Any other mixed or multiple ethnic backgrounds | 1 |
| Peer support role | Peer Worker | 23 |
|  | Senior or managerial peer support worker role | 12 |
| Peer support setting | NHS* | 21 |
|  | VCFSE** sector services, e.g. charities, community-based groups, or education settings | 14 |
| Contract type | Permanent contracts | 20 |
|  | Fixed term contracts | 15 |
|  | Self-employed | 1 |
| Employment type | Full time | 17 |
|  | Part time | 18 |
| Length of time in peer role | <1 year | 11 |
|  | 1-2 years | 9 |
|  | 2-5 years | 9 |
|  | 5-10 years | 4 |
|  | >10 years | 2 |
