## Supplementary material for "Understanding the barriers and facilitators to delivering peer support effectively in England: a qualitative interview study": Table 2

***Table 2****: Key facilitators and barriers to delivering effective peer support identified by system level*

|  | **Facilitators** | **Barriers** |
| --- | --- | --- |
| **Micro Level:** | - Having flexibility and autonomy in the role to allow delivery of person-centred care. - Having a structured workplace and job role to provide clarity about the role, support structures, and boundaries of what a PSW does and doesn’t do. - Being to some extent outside the clinical team to ensure that the PSW can act as an ally to service users. - Seeing impact of PSW for service users to provide motivation and encouragement for doing the work. - Disclosing a shared experience with service users as way to build relationships. - Having self-awareness of boundaries (e.g., what you are happy to share with service users and colleagues), including understanding of own triggers. - Having training on safety and boundaries regarding sharing personal experiences to support personal and professional working structures. | - Role ambiguity can create confusion about what PSW is, or what a PSW does/ doesn’t do. - Lack of management and support. - Having too much structure and a lack of flexibility in the role hampering delivery of person-centred care. - Being too boundaried about lived experience can hamper relationships with service users, knowledge of PSW, and can block the ‘unique element’ of what PSW does. - Lack of standardised policies about disclosure of shared experience. |
| **Meso Level:** | - Having a good working relationship and trust with present and receptive manager or supervisor. - Having a choice of options for support, supervision, and/or reflective practice, including choice for an external supervision - Flexible working conditions, allowing for reasonable adjustments for mental health needs - Having connections with other lived experience colleagues, particularly having a manager or mentor with lived experience to provide support and guidance around using lived experience professionally. - Access to learning and training, particularly training relevant to area of mental health working, e.g., specialist services such as eating disorder services. - Strong relationships with wider team members to harness a feeling of connection and community. - Professionalisation of the PSW role, including completing qualifications in the area, increases feelings of validity and value of the PSW role. | - Lack of clear and timly management, supervision and support structures. - Lack of manager or leadership buy-in for PSW reflecting a lack of value at this level placed on PSW. - Universal PSW training misses the needs of those in specialist areas of mental health (Eating Disorders, Early Intervention in Psychosis, inpatient etc). - Lack of training beyond core PSW training, e.g., statutory and mandatory training. - Variations in support depending on employer e.g., NHS vs 3^rd^ sector. - Isolation and disconnection from other peer workers, and other non-peer worker colleagues. - Assimilation/ institutionalisation into clinical teams making PSWs feel uncomfortable or losing the unique skills and elements of what peer support is. - Lack of understanding from others and self about what the PSW role is (and is not). - Professionalisation and assimilation to clinical structures of PSWs leading to a loss the nature of PSW |
| **Macro Level:** | - Investment in workforces shows systemic value for PSW - Seeing career opportunities. - PSWs in workforce plans or policies provides hope and value for future of PSWs. - Flexibility of peer support pathways and employment arrangements. - Embedded training to wider staff across and throughout system to understand the role of PSWs and create a culture change towards accepting and acknowledging the role of Lived Experience. - Having a variety of PSW roles means it can be applied across a range of settings and availably across the MH system. | - Inconsistent funding for PSW affecting job insecurity, implementation, structures for supporting the working of PSW, and longevity/ future planning for PSW within a service. - Hard to evidence impact and make a case for PSW funding. - Lack of resources e.g., computers, office space etc. - Low pay and a lack of career progression/ opportunities for PSWs leading to a culture lacking the value of PSW within and across a service. - Losing good skills/ people because of low pay/ banding/ needing to go elsewhere to progress. - Lack of equity with other professionals. - Regional inequalities in resources and funding. - Service variation and inequalities - Complexity of the MH system - Lack of service availability for ‘client’ makes work harder e.g., signposting, and makes PSWs feel hopeless - Lack of policies and plans for implementation and support for PSW more widely shows lack of value of PSW - Changing structures creates an unsteady environment for implementing PSW. |
