## Supplementary material for "Understanding the barriers and facilitators to delivering peer support effectively in England: a qualitative interview study": Supp file 2

S2 – topic guide

| **Main questions** | | **Additional Prompts** (if you need them but no need to ask them all) |
| --- | --- | --- |
| 1. | **How would you describe your role as a peer-support worker?** | - Can you walk me through an average day in your role? - How long have you been in this particular role? - Have you had any other peer support roles, including paid or unpaid roles, before this one? |
| 2. | **How have you found the role of a peer-support worker?** | How does this role live up to your expectations of being a peer support worker?  Does the job title/role and what you actually do match up?  If you have worked in other peer support roles how does this role compare?  Do you enjoy this role?   - Why/ why not? - Can you give me some examples of what you enjoy in the role?   What do you not like about the work?   - Can you give me some examples of what you find challenging in the role?   Are there any barriers that you’ve faced in your role as a PSW?   - How could these barriers be overcome to improve your experience?   Are there things that you are asked to do that you don’t feel should be part of a peer support worker role?  For you personally, how do you use your lived experience?   - Are you able to use your lived experience within your role to the extent that you’d like to? - If yes, can you give me a few examples of how you do this? - If not, can you give me examples of when you would have wanted to use LE but couldn’t? - What would help with this?   Have you had the chance to shape the role?   - Can you give me some examples of how you’ve done this? - Have any adjustments or accommodations been made to suit your needs?   If you had a good idea, do you feel you would be able to share these ideas, advocate for change or challenge the way things work in this role?   - Why/ why not? Can you give some examples of why this might be? |
| **3.** | **How would you describe how you work with people you support?** | Can you tell me a bit about the kind of people you are supporting and how you work with them?  Do you use any particular approaches, resources, or structure to your sessions or is it entirely open/flexible depending on the individual?  Do you have flexibility in how you work with the people you support?  Has how your job changed over time?  Are there any outcome measures or evaluation tools that are used in relation to your work?   - How do you feel about these?   Do you think it's generally possible to capture the impact of your work?   - If yes, how could this be done best in your opinion? - If no, why do you think not? |
| **4.** | **Are there any ways that you feel your work has had an impact on the service users you work with?** | **If yes**: can you tell me a bit more about why you feel that way   - could you share some examples of that impact?   **If no:** can you tell me a bit more about why you feel that way or what the barriers are for creating impact?  What is the biggest success you’ve experienced in your role when thinking about impact and outcomes for the service and/ or people you work with, e.g., service users? |
| **5.** | **Are there any forms of training that you’ve had for the role? If so, could you describe these?** | Did you have any training for the role/ or are you waiting for any training as part of the role?   - Is the right training for your needs? - Is it relevant to your role? - Too much/ too little? - Who provided training? - Was it online or in person? - What aspects of the training did you find useful/ not so useful? - Have you had any training around how to use your lived experience?   Is there any training you would have liked to have had?  What would you include in a training package for a new peer supporter |
| **6.** | **W*hat opportunities, if any, do you have for supervision/reflective practice?***  Definition: "A specialised form of mentoring provided for practitioners responsible for undertaking challenging work with people. Supervision is provided to ensure standards, enhance quality, advance learning, stimulate creativity, and support the sustainability and resilience of the work being undertaken."  OR  “In health and social care, supervision involves the regular meeting of a manager or senior practitioner with staff members, with the aim of reviewing and reflecting on performance, providing support and improving practice.” | **If Yes:**  What is supervision like for you?  Can you talk me through what it looks like and perhaps give some examples of how this works and influences your work - *there might be multiple sources of supervision e.g., line management from one person, supervision or mentoring from another – do you have opportunities to take into these different supports? (line-manager; peer-group, someone outside of your organisation)*   - Who provides supervision? - What’s discussed? - Who sets agenda? - How often do you have supervision? Is this group or 1:1? - How do you feel about the amount of supervision that you get? - Do you get support from anyone with lived experience – formally or informally?   **If no:**  Do you know why not?  Do you want supervision?   - If yes: Can you talk me through what this would ideally look like for you and give some examples of how it might work in practice? - If no: why not? |
| **Break? – We’ve reached about halfway so it might be the perfect time to have a quick break and grab a tea, have a walk about etc…** | | |
| **7.** | **Could you tell me about your working relationships with colleagues?** | How would you define ‘your team’, e.g., what type of colleagues do you work alongside, clinicians, academics, other peer workers?   - How do you work with your team? - Can you give me some examples about how you work with different people in the team? - How much do you feel you are part of the team? Why/ why not?   How do you think you/ lived experience roles are ‘seen’ within the organisation?   - Can you give me some examples of why you feel like that?   Do you feel your role/ lived experience is valued by your team?   - Why/ why not? Can you give me some examples of why you feel this? - Do you feel that your opinion is listened to? How do you know that’s the case? |
| **8.** | **We are going to talk more widely now about how your role fits in the organisation as a whole. How do you see peer support within this wider structural perspective?** | Do you feel your role is valued in the organisation?   - Why/ why not, - Can you give any examples to help me understand this better?   What other lived experience roles are there within your organisation?  Is there any organisation-wide strategy for lived experience roles?  Has there been any scope to influence this by you or other lived experience practitioners within your organisation?  What organisational changes would you like to see to improve peer-support work, if any?  What values do you think organisations need to make peer-support work well?  Do you think there are there any organisational factors that influence whether or not teams get peer-support workers?  Does your role work in partnership with other organisations – if so, what has been your experience of this? |
| **9.** | **Please tell me about any ways in which national policy initiatives or guidelines influence your work?** | Are there any recent changes in mental health services that have had an impact on peer support roles from your perspective?  Are there any key policies (e.g., the community mental health framework) that have had an impact (positive or negative) on peer support from your perspective? |
| **10.** | **How do you see the future of peer support work?** (Biggest concern? Biggest hope?) | What is you biggest concerns?  What are you biggest hopes?   - How could these be achieved? Examples?   What would you take away message to policy makers about peer support work in England? |
| **11.** | **Is there anything else you would like to share with us about your experience as a peer-support worker?** |  |
