## Supplementary material for "Understanding the barriers and facilitators to delivering peer support effectively in England: a qualitative interview study": Supp file 3

Supplementary table: S2

| **Theme** | **Subtheme** | **Summary** | **Illustrative Quotes** |
| --- | --- | --- | --- |
| **Micro level: doing the work** | **‘You need flexibility and structure in this role’: striking the balance** | The importance of flexibility in meeting people where they are. | *I would describe it as very flexible. It’s very personal, it depends on the individual and what they want out of peer support. Sometimes they just want someone to talk to with similar experiences. Other times they want more practical help... I think compared to clinical roles, it’s a bit less structured. I have more freedom over how many sessions I can have with a person. it depends on each person. (PT01, NHS)* |
|  |  |  | *I’ve had so much freedom in the role to develop it myself, being the first peer support worker in our team. ... I’ve been able to take initiative. I’ve learnt a lot. My team trusted me so that has been really positive (PT07, NHS)* |
|  |  | The challenge of being too flexible and unstructured, leading to the feeling that the role becomes unboundaried. | *I think it’s a very flexible role. But then sometimes that can be a downside. When I first started, because there was so much freedom, I didn’t know where to start. In the beginning, I’d go into a session and not really know what to do. But I feel like I’ve got a better grasp now. But in the beginning, it was a little bit difficult. (PT01, 3^rd^ sector)* |
|  |  |  | *The role of peer support worker isn't really defined, and we find ourselves doing things that maybe aren’t within the stuff that we should be doing… (PT01, 3^rd^ sector)* |
|  |  |  | *I’ve been asked to make phone calls to certain people or make contact with certain people and it’s not necessarily peer support as I understand it. Perhaps my boundaries aren’t as good as they should be… I’m finding myself doing a lot of housing support and trying to get in contact with the council and this and that. (PT05, NHS)* |
|  |  | The challenge of having too much structure. | *The peer support worker role was being pushed more into clinical-like roles, so the heart of peer support was being lost a little bit, in the midst of more ‘admin’y’ stuff and more paperwork and more notes. (PT01, NHS)* |
|  |  | Concerns that the role was becoming too closely integrated into standard clinical practice. | *(Currently we've got new managers coming in who want it set out ... a bit more clinical. They want a menu system of what I offer. I don't know. For me, that doesn't- I struggle with that, a bit... And I feel like if I'm offering menus I lose a bit of that person-centredness, perhaps. (PT12, NHS)* |
|  | **"We work differently": having a shared connection with service users’** | A shared experience facilitating the connection between PSW and service user, which goes beyond how clinicians can. | *We work differently. I think the biggest difference is that we immediately have that empathy… by being a veteran with lived experience I automatically have that empathy, because I've been in their shoes, and they realise that quite quickly... And conveying that over to people I think is the most powerful thing that we can do in peer support. (PT14, 3^rd^ sector)* |
|  |  |  | *I heard this term/phrase said a lot, and it’s, “Peer support workers can say the things that we can't say.” Psychologists/therapists, ... I think they feel confined in that clinical role, whereas we can be more open, I think. Yes, that’s what they’re wanting us to do. (PT32, NHS)* |
|  |  | The shared experience as a means to reducing the power dynamics and hierarchies that exist within healthcare services. | *Within my work, I've been the only black male... so I think I'm in a unique position to share my experiences. ... [Professionals] they'll come to me with experiences where they might be working with someone that is black or from an ethnic minority, and they might ask me questions of how I would deal with it, or how would they approach a certain way, or what culturally would they need to be aware of? Or is there a religious thing that they need to be aware of, or certain things like that? So I guess I can help in those respects. (PT34, NHS)* |
|  |  | Intersection of shared experiences as a facilitator to connecting with service users from similar backgrounds, but also as a barrier due to stigma, discrimination and othering. | *[A service user] came in, was really quiet and really low, and then all of a sudden, was very explosive and just really, really unhappy… the way that we did connect was that he kind of said, “Well, what’s your experience of mental health problems anyway?” … It’s one of the principles of being a peer, is that he does understand that I’ve been through something similar and that I’m not just someone who’s trying to move him along. (PT05, NHS)* |
|  |  |  | *Being someone from an ethnic minority background, you go into a situation sometimes where you might be being judged or they [the client] might be a bit hesitant to work with you… there might be some comments or there might be some discrimination. In my team, there are not that many people from ethnic minority backgrounds. So that can be a barrier for me sometimes, but it's something that you can break down a lot of times with people and get them to be on your side. (PT34, NHS)* |
|  | **‘It’s important to be open about my experience but you have to have boundaries’: being open but boundaried** | The importance of using boundaries to keep the work healthy and safe for both the PSW and SU. | *I only talk about my experience if I think it's in a hopeful way and it’s going to be supportive to the person. It’s not going to be harrowing for them to hear, if you know what I mean, it’s not easy listening to people’s stories. (PT07, NHS)* |
|  |  |  | *With eating disorders, there’s a lot of desperation and people will just ask me really direct questions, like, “How did you recover? How long were you ill for? What can I do? What did you do about your confidence?” They're really interested in learning what our experiences were. And, obviously, we have to be super boundaried with that, but I know in other peer support roles it seemed more about them and you’re there as a safe person, because of that lived experience. But it seems like people in the eating disorder service are really actually wanting advice from someone from lived experience, which is difficult… it does feel really different to other peer support roles. (PT32, NHS)* |
|  |  |  | *With the peer support, I can only go as deep as I can, because I am not paid to be a counsellor... I have experienced burnout working so I need to keep those boundaries in and set, so that I can carry on with the work, because it is, literally, emotionally tiring. People will not realise that. (PT28, 3^rd^ sector)* |
|  |  | The role of training to facilitate boundary setting. | *I decided on the boundaries that I set now. And that training also helped me to reflect upon how I could go about saying these things. (PT34, NHS)* |
|  |  | Having self-awareness of their own triggers, mental health and recovery journey were seen as crucial in creating boundaries that kept them safe in a peer role. | *You need to know that you’ve got the ability to speak up when you’re not okay. If you can’t even speak up when you’re not okay, in this role you’re going to really struggle. You need to be able to have the awareness of yourself. You need to know what you’re going to do if that happens. (PT09, NHS)* |
|  |  |  | *The important thing to learn in peer support is self-care. “If you can’t look after yourself then you can’t look after anybody. So, if you’re affected by somebody else’s trauma then you need to raise your hands and come and speak to us and go, ‘Look, I've got an issue here/there’, or whatever.” (PT14)* |
|  |  | Boundaries were more easily upheld when there is a clear job role, expectations and limitations. | *With the peer, for the safety of the participants, I really limit myself to a particular level. I do not go as deep as possible, because aside from the group support, they’re also receiving some form of mental health support from the NHS. So, I leave it to their own counsellors to go there, not me. (PT28, 3^rd^ sector)* |
| **Meso level: working within the organisation** | **‘Sometimes peer support is really, really hard…’: the importance of good support and supervision** | The importance of good support to manage the emotional impact the role can have., contracted with the lack of support where this is less available. | *[I] feel really supported… “Do you know what, sometimes peer support is really, really hard.” Whatever the concern is I can go to [my manager] and talk through stuff. I feel really supported. Those are my formal supervisions, but if I needed to speak to [my manager] today, for example, they’d respond and find time for me. I’m really lucky in that aspect. (PT05, NHS)* |
|  |  |  | *We don't currently have it [consistent supervision or reflective spaces]. But it is something that we're trying to do… There's a lot more support for that in the NHS. There's a lot more structure. There's processes to support, and there's a lot more training. And there isn't that [here] - it's down to resources, I think, a lot of the time. In the NHS, you have got more resources. Whereas in the charity sector, you don't. (PT21, 3^rd^sector)* |
|  |  | Having flexible support from a range of structures, from different team members, and with different elements was beneficial to meeting the changing needs of PSWs. | *They bring stuff. I bring stuff. We discussed what the layout would be. I’ve always said I want to talk about wellbeing first, before we get into anything else. We've only recently started taking on clients, so before it was mainly just talking about admin stuff, but hopefully now there’ll be more of a client focus so I can actually get support with that. (PT32, NHS)* |
|  |  |  | *Sometimes if I felt it was more relevant to talk to one of the clinicians or the doctors... I would ask them, “Is there any chance you've got a spare hour this month or this week that I can check in with you about something?” Mostly, they're happy to oblige if they've got a bit of free time. (PT06, NHS)* |
|  |  |  | *So, I just learn so much by being around him. You know, he’s not technically my line manager, but he’s probably been the most invaluable source for me because just hearing someone do their job who’s so experienced is really, really helpful. Then I also get to ask questions whenever I have them, which I feel very lucky about. (PT05, NHS)* |
|  | **‘No peer is an island and all that’: the impact of the wider team** | Having strong relationships with members of their team facilitated PSWs to do their job, and having trust in colleagues was central to forming a positive connection. | *My team that are close to me, I’ll speak to them once a week for an hour again on top of the supervision. Again, that is probably more like bringing work problems together and trying to solve them, rather than me talking about my peer support experience… this one is just for me and my two team members to talk about the project, my problems within that rather than other problems. (PT04, NHS)* |
|  |  |  | *I am considered a very vital part of the team, regardless of me being a peer support worker. ... Nurses and the doctors will always ask about how I feel about something or what I would consider. If I've got an idea, I'll share it, and I get listened to, and it gets put in their plan, their care plans. So, they do, and they definitely appreciate it, because again I know because it's not just me giving a suggestion and it falling on deaf ears. (PT06, NHS)* |
|  |  | Connection with other PSWs or lived experience workers who had a shared experience of doing the job was felt in facilitate PSWs in their roles. | *A lot of my colleagues have lived experience, I think it’s really positive. We meet once a week just to chat for half an hour and then I have another colleague who does a very similar role to me... and we’ve started meeting, kind of, using each other as a sounding board, I think. If we have a difficult conversation with someone, we’ll chat it through afterwards. (PT18, 3^rd^ sector)* |
|  |  |  | *[A Peer worker] started at the adult team, they were so lonely they got linked in with me just for an hour to say hello and how’s peer support going. She literally cried. She was like, “I’m so lonely, no one knows what to do with me.” She was coming across so many problems. (PT08)* |
|  |  | Colleagues’ assumptions and fears regarding individual’s vulnerability or risks of PSW. | *I am aware there’s been concerns of risk and worrying… clinicians having that concern of us [peer support workers with lived experience] being in the service … the team was so welcoming when we joined but some people weren’t hesitant but were concerned. (PT09, NHS)* |
|  | **“Do you know what peer support is?”: the heterogeneous nature of the peer support role** | The heterogeneous nature of PSW lead to role ambiguity and lack of understanding about the role both personally and within wider team members. | *Its’ been funny that I have to just ask, “Do you know what it is?… I think it would be good if there was a bit more sort of awareness and education for other people that aren’t peer support workers, about what peer support is, because like I said, a lot of people, we go in and they don’t really know what a peer support worker does and they don’t know what our roles are. ” (PT05, NHS)* |
|  |  |  | *It’s quite new in our trust. I don’t know whether there’s so much awareness of it. When I’m training with colleagues from [elsewhere], it’s just a massive difference. I just hope one day that maybe our trust will get to where some of the [other] trusts are. I just think it’s amazing what they’re doing with all the career progression and opportunities for peer workers to progress there. (PT07, NHS)* |
|  |  |  | *[Clinicians have said] "We haven't got anything else to offer. So, we'll offer peer support." And I don't necessarily think that there's always an appreciation of what peer support could bring to the service. (PT21, 3^rd^ sector)* |
|  |  | Role ambiguity was more pronounced in services where peer support work was new in the organisation, compared to where peer support was established. | *I think because there are strategies and stuff in place in the NHS, we are more established. And the role is definitely more established, it's very clear what a peer support worker does… Whereas in the third sector, because there isn't a strategy. I know that sometimes my job title gets forgotten. I have been to meetings where I have just been introduced as a good life facilitator. And obviously, it's not the end of the world. But when you're not established like you are in the NHS, you don't have the processes in place, it can just all smudge into one similar colleague group. Which is not great. (PT21, 3^rd^ sector)* |
|  |  | Managerial and leadership support and buy-in to PSW was felt to facilitate the development of such policies and strategies to embed these roles. | *I think it's who really is running those teams and what their values or beliefs are, and the value of peer support in their minds, whether they push for it to be funded. Like I say, my new manager, he'd have three more of us, and he's asked for funding. And there's a community team based in another building on the premises who have got funding and won't employ a peer support worker. (PT12, NHS)* |
|  |  | The increased 'professionalisation’ of PSW as a role as both a facilitator to having peer support valued by others where it has previously been overlooked, but also as a barrier by restricting the flexibility needed to do the role. | *I know that sometimes people can be a little bit wary of a peer support worker, like, "Okay, but they haven't been to uni. They haven't studied. They haven't done everything else that everyone else has done." (PT12, NHS)* |
|  |  |  | *There have been a couple of people who, in the beginning, didn’t want to interact with me, because I wasn’t a professional. (PT30, NHS)* |
|  |  |  | *A negative I think is that it becomes like the whole professionalism of the NHS… that the peer support role will be seen in a similar way… And it takes away the informal warm friendly vibe... we’re not like a healthcare assistant, and I think sometimes it can be viewed like that in the NHS a bit. (PT18, 3^rd^ sector)* |
| **Macro level: beyond the workplace – systemic factors** | **‘The frustration of it is not having funding for peer support workers’: the impact of limited funding and resources** | Restrictions on the role due to funding limitations faced by services that have a direct impact of the PSW role. | *Having done it for a year there are frustrations I think because it’s such enriching work and because it’s so important, the frustrations of it not being there or not having the funding to have more, not to get further training and not to upskill us is really hard to stick with. It feels frustrating for the patients as well. Working alongside people who want more, but you can go, “Sorry, I can only do these six sessions because that’s all I’m allowed to do. I need to see somebody else because there’s not enough of us.” More of us please. (PT08, NHS)* |
|  |  | Barriers created due to restrictions on resources that are funded for doing the day-to-day aspects of the role. | *Another thing is I’ve been doing the role for three months and I still don’t have a work mobile phone…. There are lots of sort of tech issues. So it feels like we need a laptop, but yes, for some reason, we’re the only people in our service who it’s, for some reason, up for debate as to whether we need a laptop or not… we’ve had new people come into the service who are support workers or community connectors and they’ll get a phone right away. (PT05, NHS)* |
|  |  | The implication that lack of funding is related to the lack of value placed on PSW. | *The biggest worry is funding or somebody higher up somewhere, somebody somewhere going, “This isn’t important, we don’t need it. They’re not clinicians, let’s scrap it.” I think that’s really scary to think peer support wouldn’t be here. (PT08, NHS)* |
|  | **‘There are a lot of problems around retention, people leave because they need money!’: low pay and lack of progression** | For some people low pay was felt to represent the lack of value placed on PSW in the mental health system, however this wasn’t a universal issue showing the different spaces that PSW hold for different people. | *I think the peer support workers at lower pay grades are over-utilised and under-appreciated for their value for money. (PT14, 3^rd^ sector)* |
|  |  |  | *It might sound corny, but money hasn't really been an issue because I've never had money. I've always been not working, or under so much debt, that we could never get credit. So, we've had to get by without money. So, now, I feel like I'm a millionaire. Because that is just unbelievable. So, I feel I'm well-paid. Yes, I just love it. As I say, I volunteered, and I loved that, and I would have been quite happy doing this voluntarily. But the fact that they want to pay me, that's- I'm happy with that. (PT12, NHS)* |
|  |  |  | *I'll just tell you at the Trust, they do one to one sessions and they do groups as well. So, they do both and people are paid different to others. It's just madness. It's just like, “What are you doing?” (PT03, NHS & 3^rd^ sector)* |
|  |  | The barrier that a lack of career progression creates for PSWs who see the limited development they face in this area, which doesn’t reflect landscape for colleagues in nursing or psychology. | *I think I’m concerned about the lack of development for peer support workers. We come in and then what, this is our career forever. Not necessarily, but where’s the growth for us? We have aspirations too. (PT08, NHS)* |
|  |  |  | *There's a massive thing about peer support progression because once you've become say, in the NHS, a senior peer support worker, where do you go from that? The next step, they say, at the moment, is to become a social worker, or a nurse, or stuff like that. But actually, then you lose your lived experience element. You lose what actually you want to be. But at [service name], there's nothing at the moment. There isn't even like a senior role. So, if I wanted to move up, I would lose my lived experience peer support, peer specialist title. And I think that's a barrier to a lot of people. Like people do just go into nursing, or whatever, and forget about their peer support core. Because there isn't the progression. (PT21, 3^rd^ sector)* |
|  |  |  | *“I'm really happy in my role. I don't want to be a nurse.” (PT06, NHS)* |
|  | **‘It’s really complicated!”: Working in complex mental health systems** | The complexity of working in systems that have their own politics and structures which create barriers for PSWs. | *I’ve worked for the NHS since I was 19 so I understand the policies, the processes, the procedures. One of our colleagues hadn’t worked for the NHS before and found it overwhelming. Just the fact that trying to get hold of IT is a massive palaver, it’s not simple. When you’re somebody that’s got lived experience all of that can be far too much to process. (PT09, NHS)* |
|  |  | Being held back by barriers within the systems that are beyond the control of the PSW. | *Sometimes this person, for example, is too complex for IAPT because they’ve mentioned suicide. Then they’ll come into our referral meeting and they’re not complex enough for us. And then psychology won’t take them off because they’re maybe not in a stable place to do trauma-informed therapy…. I think that can be a little bit challenging, and I don’t really think it’s anyone’s fault., it’s just a difficulty with the system… there’s a bit of that like it doesn’t feel like we’re always working in a recovery-oriented way. (PT05, NHS)* |
|  |  |  | *Going back to the geographical characteristics of our patch... it’s predominantly rural. It’s really big. So, that puts constraints [in what can be done] because there’s only me, I cover the whole area. (PT07, NHS)* |
